# Smartphone-based measures as real-world indicators of functional status in advanced cancer patients

**DOI:** 10.1101/2025.02.04.25321686

**Authors:** Marcin Straczkiewicz, Nancy L. Keating, Stephanie M. Schonholz, Ursula A. Matulonis, Neil Horowitz, Susana Campos, Jukka-Pekka Onnela, Alexi A Wright

## Abstract

**Objective:** This study evaluated the feasibility of using smartphone-based metrics to monitor physical functioning and quality of life in patients with advanced gynecological cancers. We analyzed associations between gait (step count, cadence, stride acceleration) and measures of mobility (home time, distance traveled, number of significant locations visited) with patient-reported outcomes (PROMs).

**Methods:** We studied raw accelerometer and GPS data from smartphones over 180-days for 85 patients with advanced gynecologic cancers. We computed measures of gait and mobility from smartphone sensor data, and PROMs (performance status, health-related quality of life, physical functioning) from smartphone surveys at baseline, 30, 90, and 180 days. We assessed longitudinal associations of digital gait and mobility measures with PROMs using linear mixed-effects models, with attention to smartphone adherence and temporal trends.

**Results:** Smartphone adherence was high: 83% of participants reported daily smartphone usage >1 hour; 74.1% had >16 hours of daily use. Gait measures, particularly step count and stride acceleration, were statistically associated with PROMs. Worsening patient-reported ECOG performance status corresponded to reduced step count (ECOG 3 vs. 0: -1837 steps/day, *p*<0.001), while higher PROMIS Physical Function was associated with step count increase (+72.64 steps/day per one-point increase, *p*<0.001). Mobility measures were less strongly associated with PROMs but provided complementary insights into patients’ behavioral patterns.

**Conclusion:** Smartphone-based gait and mobility metrics offer robust, real-world insights into individuals’ health statuses, providing a scalable, low-burden alternative to wearable devices. The high levels of smartphone use among participants underscore the feasibility of integrating this technology into routine oncology care.

## 1. INTRODUCTION

More than half of patients with advanced cancers experience serious symptoms that interfere with daily living and erode patients’ quality of life[1]. In clinical trials, remote monitoring of symptoms using patient-reported outcome measures (PROMs) improves cancer patients’ quality of life[2,3], physical function[4], and survival[5,6]. However, real-world implementation of remote monitoring of PROMs is intensive[7–9], dependent upon patient adherence and clinician responsiveness[9,10], and may further exacerbate existing disparities[9,11].

The use of sensor-based digital health technologies (DHTs), such as commercial wearables, represents a promising strategy for obtaining objective measures of advanced cancer patients’ functional status and has been shown to predict hospitalizations and death in small studies[12–14]. However, commercial wearables are expensive and critically dependent upon patient adherence. Smartphones may represent a scalable alternative for remote monitoring of patient physical functioning and symptoms without additional cost or burden. To date, however, clinical validation of smartphone-based measures of patient activity has been limited, especially over longer time periods[15,16].

To address this gap, we examined whether raw smartphone accelerometer and Global Positioning System (GPS) data, collected from participants’ personal smartphones, could enable the remote monitoring of advanced cancer patients’ physical functioning and quality of life. We examined three gait-related measures of physical activity derived from accelerometer data: step count, gait cadence, and stride acceleration; and three measures of mobility from GPS data: home time, distance traveled, and number of significant locations visited in a 24-hour period[17–20].

In this study we aimed to examine: 1) associations between digital measures of physical activity and mobility and three PROMs: functional status, health-related quality of life, and physical function; 2) longitudinal changes in digital measures over time; and 3) the association of reported smartphone use and adherence with data quality.

## 2. METHODS

### 2.1. Study Design

We conducted a pilot study to assess the feasibility, acceptability and perceived effectiveness of a mobile health intervention that used commercial wearables and the Beiwe platform[21] to collect passive data on patients’ behavior using their personal smartphones. Briefly, patients with recurrent gynecologic cancers receiving systemic therapy (i.e., chemotherapy, immunotherapy, anti-angiogenic, or targeted therapies) were recruited from the Dana-Farber Cancer Institute in Boston, MA[22]. During a pre-pilot (n=10), participants received commercial wearables, installed Beiwe on their smartphones, completed electronic patient-reported outcome measures (ePROMs, not included in this study) on their smartphone, and completed PROMs at baseline and 30 days[22]. During the pilot randomized controlled trial (RCT), participants (n=75) were randomized in a 2x2 factorial design to receive: 1) a commercial wearable, 2) ePROMs, 3) a commercial wearable and ePROMs, or 4) neither. Participants in the pilot RCT completed PROMs as baseline, 30, 90, and 180 days. All participants contributed passive sensor data (accelerometer and GPS) using Beiwe and all participants provided written informed consent. The study was approved by the Dana-Farber/Harvard Cancer Center Institutional Review Board and was registered in clincialtrials.gov (NCT03022032).

### 2.2. Patient-reported outcome measures (PROMs)

Functional status: Participants reported their performance status using a patient-adapted version of the Eastern Cooperative Oncology Group Performance Status (ECOG PS)[23], which has been shown to predict ED visits, hospitalizations, and survival in patients with advanced cancers[24–26]. The score ranges from 0 to 4, where 0 is defined as: “I am fully active, and able to carry out activities the same as before my cancer diagnosis, without any restrictions,” and 4 is defined as: “I am completely disabled, cannot carry on any self-care, and am totally confined to a bed or chair.”

Health-related quality of life: Participants completed the EQ-5D-5L, a well-validated 5-item questionnaire that assesses mobility, self-care, usual activities, pain/discomfort, and anxiety/depression[27]. The items are scored on a scale of 1 to 5, with lower scores indicating better status.

Physical function: The PROMIS Physical function (PF) 6b is a brief measure of patient health status that is not disease specific[28,29] but has been validated in patients with advanced cancer[30]. It consists of six questions assessing patients’ overall health, level of physical disability, and general well-being. The items are scored on a Likert scale of 1 to 5, with higher scores indicating better health.

At baseline, participants also estimated their smartphone use in response to the question: “Over the course of a day, approximately how much time do you spend using your smartphone?” Response options were: “More than 4 hours,” “Between 2 to 4 hours,” “Between 1 to 2 hours,” “Between 30 minutes and 1 hour,” and “Less than 30 minutes.”

### 2.3. Digital measures

At baseline, participants installed the Beiwe application to collect raw accelerometer data in a three-dimensional orthogonal coordinate system with the default sampling rate (10 Hz or more) and alternating sampling scheme (10 seconds of data collection followed by a 20-second pause) to prevent draining participants’ smartphone batteries[31]. Beiwe also collected GPS data for 1 minute followed by a 10-minute pause. The missing GPS data were imputed using methodology based on sparse online Gaussian processes published previously[20]. The sensor sampling scheme was identical on all participants’ smartphones. Pre-pilot participants were followed for approximately 30 days, while the participants in the pilot RCT were followed for approximately 180 days.

We examined three gait-related measures of physical activity derived from raw sub-second level smartphone accelerometer data: step count, gait cadence, and stride acceleration.

Step count was estimated as a sum of one-second step frequencies identified using a previously developed and validated open-source method[17,18]. Gait cadence was estimated as an average step frequency per minute from all periods of physical activity classified as walking by the method. Stride acceleration was estimated as an average acceleration collected during automatically segmented and rescaled gait strides (two consecutive steps) and corresponds to an individuals’ gait intensity[32].

We also examined three mobility measures from imputed GPS mobility trajectories: time spent at home, distance traveled, and the number of significant locations visited in a 24-hour period. Home time was estimated as time spent in the most frequently visited location for a person between 8pm and 8am each day over the course of the data collection. Distance traveled was estimated as a sum of trip lengths between two locations without a directional change or pause. Significant locations were distinguished as distinct pauses at least 15-minutes long and 50 meters apart[20].

### 2.4. Statistical analysis

Digital measures were summarized on a daily level. Participants’ step counts were adjusted proportionally to the on-off sampling scheme of data collection. All analyses were conducted using observations from days with at least 960 minutes (16 hours) of collected data, henceforth called “valid days”[33]. Means and standard deviations (SD) were used to summarize the data. To align gait and mobility measures with PROMs, we computed an average of participants’ daily digital measures within ±10 days from each completed survey.

We examined the quality of smartphone data collection by examining the proportion of 1) the number of actual vs expected days of smartphone data collection and 2) the number of actual vs. expected “valid days.” We used linear mixed models to determine associations between participant-reported smartphone use (reference category: “more than 4 hours a day”), the quality of smartphone sensor data collected, and the digital measures based on that data, averaged for each participant within the first two weeks of data collection. In each model, we used digital gait and mobility measures as the outcomes and survey responses as predictors.

Participants’ ECOG PS scores ranged between 0 and 4, EQ-5D-5L scores between 5 and 25, and PROMIS PF 6b scores between 6 and 30. Before analysis, participants’ PROMIS PF 6b scores were converted to T-scores[34]. We fit linear mixed-effect models to assess associations between the outcomes of interest, gait and mobility measures, and categorical (i.e., ECOG PS score and EQ-5D-5L subscales) and continuous (PROMIS PF 6b) survey responses. These models contained a random intercept and slope for each patient to account for repeated measures within patients. Similar linear mixed models with random intercept and slope were used to estimate baseline and monthly-level changes in gait and mobility measures.

Gait measures were computed in MATLAB (R2022a; MathWorks, Natick, MA)[35]. Mobility measures were computed in Python (Python Software Foundation: version 3.9.14)[36]. Statistical analysis was performed using R (version 4.1.2; The R Project). When appropriate, we report 95% confidence intervals (CI) and p-values (alpha=0.05).

## 3. RESULTS

### 3.1. Cohort summary

Data collection took place between April 2017 and December 2019 and included 85 individuals with advanced gynecological cancers, including ovarian (n=64), endometrial (n=12), and cervical, vaginal, or vulvar (n=9) cancers (**Table 1**). Participants’ ages ranged between 24 and 79 years of age (mean=61.6, SD=10.3), 78.8% self-identified as White, 8.2% as Black, 4.7% as Asian, and 8.3% as a different race; 97.6% were non-Hispanic. Participants were predominantly married or had a partner (63.5%), held a college degree (61.2%), and were not employed (63.5%).

**Table 1.** Baseline demographic, clinical, and smartphone characteristics.

| **Demographic characteristics** |  |
| --- | --- |
| Female, N (%) | 85 (100) |
| Age, years, mean (SD), median [min, max] | 61.6 (10.3), 62 [24, 79] |
| Race, N (%) |  |
| White | 67 (78.8) |
| Black | 7 (8.2) |
| Asian | 4 (4.7) |
| Native American | 1 (1.2) |
| Other | 6 (7.1) |
| Ethnicity, N (%) |  |
| Hispanic | 2 (2.4) |
| Non-Hispanic | 83 (97.6) |
| Marital status, N (%) |  |
| Married or partnered | 54 (63.5) |
| Divorced or separated | 12 (14.1) |
| Widowed | 6 (7.1) |
| Single | 12 (14.1) |
| NA | 1 (1.2) |
| Educational status, N (%) |  |
| High school or less | 11 (12.9) |
| Associate's degree or some college | 22 (25.9) |
| Bachelor's degree | 23 (27.1) |
| Graduate degree | 29 (34.1) |
| Employment status, N (%) |  |
| Full-time | 15 (17.6) |
| Part-time | 11 (12.9) |
| Not working | 54 (63.5) |
| NA | 5 (5.9) |
| **Clinical characteristics** |  |
| Cancer type, N (%) |  |
| Ovarian | 64 (75.3) |
| Endometrial | 12 (14.1) |
| Cervical, vaginal, or vulva | 9 (10.6) |
| Anthropometric characteristics, mean (SD), median [min, max] |  |
| Height, cm | 160.3 (6.3), 160 [146, 182] |
| Weight, kg | 73.6 (16.6), 70 [48, 119] |
| BMI, kg/m<sup>2</sup> | 28.6 (6.5), 28 [19, 51] |
| Smartphone operating system, N (%) |  |
| iOS | 65 (76.5) |
| Android | 20 (23.5) |

The heights of participants ranged between 146 and 182 cm (mean=160.3, SD=6.3), weights ranged between 48 and 119 kg (mean=73.68, SD=16.6), and BMIs ranged between 19 and 51 kg/m^2^ (mean=28.6, SD=6.5).

Participants’ ECOG PS, EQ-5D-5L, PROMIS PF 6b scores, and smartphone daily use are displayed in **Figure 1**. For example, at baseline, 31.8% of participants reported an ECOG PS=0, 54.1% ECOG PS=1, 9.4% ECOG PS=2, and 4.7% ECOG PS=3 or 4. Participants’ PROMIS PF 6b mean T-score was 44.4 (SD=7.4) at baseline, 43.4 (SD=8.0) at day 30, 45.1 (SD=8.2) at day 90, and 44.2 (SD=9.5) at day 180, consistent with United States population averages for patients with advanced cancer[30].

**Figure 1.**
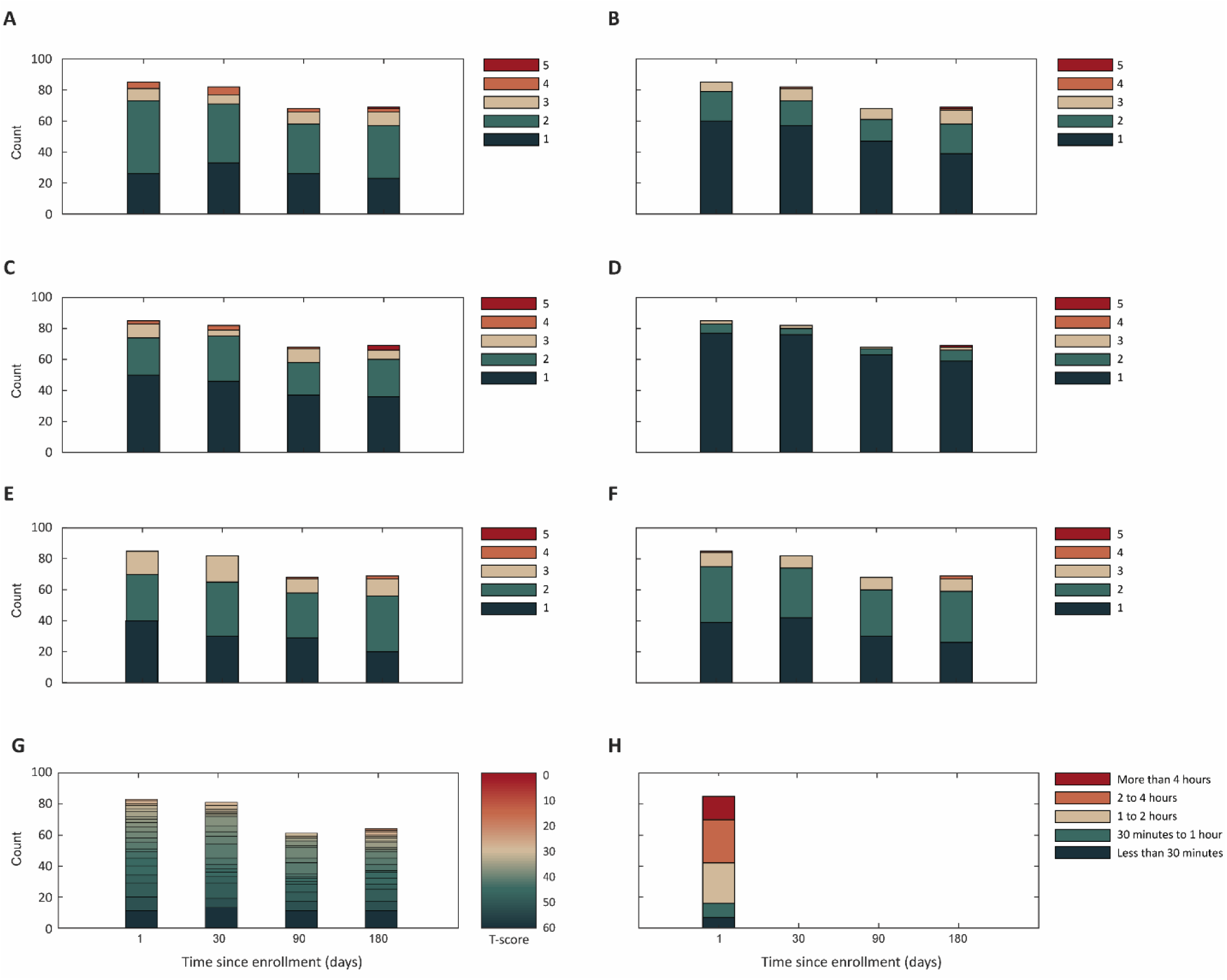
Summary of collected survey responses. In the study, we collected ECOG PS (A), EQ-5D-5L, including responses on mobility (B), usual activity (C), self-care (D), pain (E), and anxiety (F), as well as PROMIS PF 6b (G) and smartphone daily usage preference (H). Smartphone daily usage preference was collected at baseline. Functional status responses were collected at baseline, after 30, 90, and 180 days.

Study participants reported substantial smartphone use: 15/85 (17.7%) reported more than 4 hours per day, 28/85 (32.9%) reported between 2 to 4 hours, 26/85 (30.6%) reported between 1 to 2 hours, 9/85 (10.6%) reported between 30 minutes and 1 hour, and 7/85 (8.2%) reported fewer than 30 minutes per day.

### 3.2. Smartphone data quality and impact of participant-reported smartphone use on digital measures

Participants in the pre-pilot study contributed a mean of 25.1 days (SD=9.3) of smartphone sensor data, while those in the pilot RCT contributed a mean of 132.4 days (SD=54.8). After restricting sensor data to valid days (>16 hours of data/day), about three quarters of participants (63/85, 74.1%) had data of sufficient quality. This resulted in a mean of 21.1 days (SD=7.4) for the pre-pilot and 75.6 days (SD=48.4) for the pilot RCT.

In unadjusted analyses, there were no differences in the quality of smartphone data collected by participants’ reported smartphone use. Specifically, participants who reported using their smartphone for fewer than 30 minutes per day had similar proportions of smartphone sensor data as those reporting more than 4 hours per day and similar proportions of days with valid data (**Figure 2, panels A and B**). Similarly, we did not see statistically significant differences in participants’ digital measures of physical activity and mobility by reported smartphone use (**Figure 2, panels C, D, and E and F, G, and H,** respectively).

**Figure 2.**
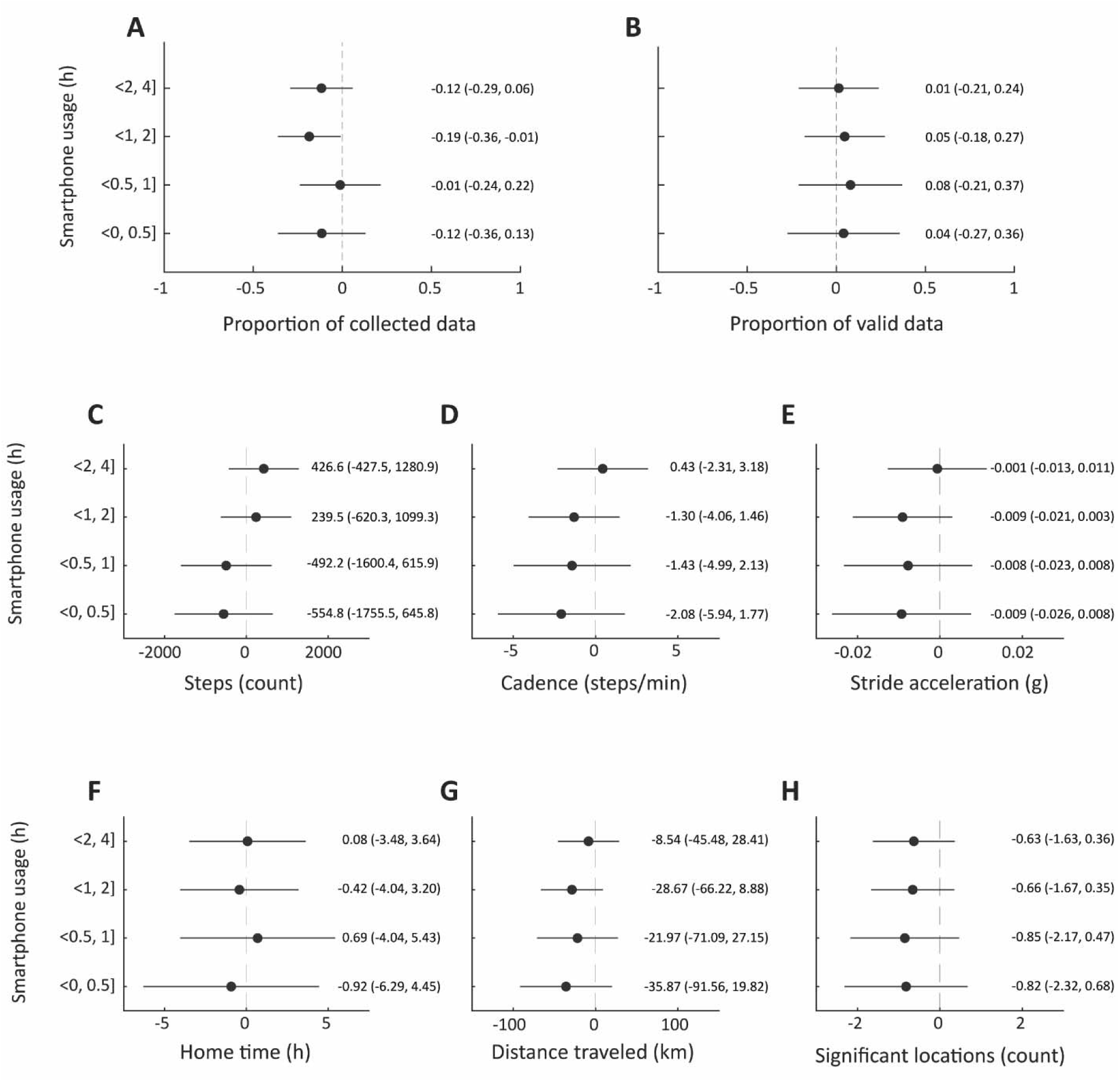
The association between patient-reported hours of daily smartphone usage and data quality (proportion of collected data (A) and valid data (B)) and digital measures of physical activity (step counts (C), cadence (D), stride acceleration (E)) and mobility (home time (F), distance traveled (G), and number of significant locations (H)).

### 3.3. Associations between PROMs and digital measures of participants’ physical activity and mobility

Figure 3 presents the regression coefficients of six smartphone-based measures of physical activity and mobility (steps, cadence, stride amplitude, home time, distance traveled, and signification locations) by ECOG PS and EQ-5D-5L subscales. As shown in **Figure 3, panel A**, smartphone-based measures of physical activity were statistically significant and negatively associated with participants’ ECOG PS such that participants with better ECOG performance status had higher step counts and greater stride acceleration, compared with participants with worse ECOG PS. Similarly, participants with better performance status were more likely to visit multiple significant locations per day, compared with worse performance status, although we did not observe any statistically significant associations between ECOG PS and time spent at home or distance traveled.

**Figure 3.**
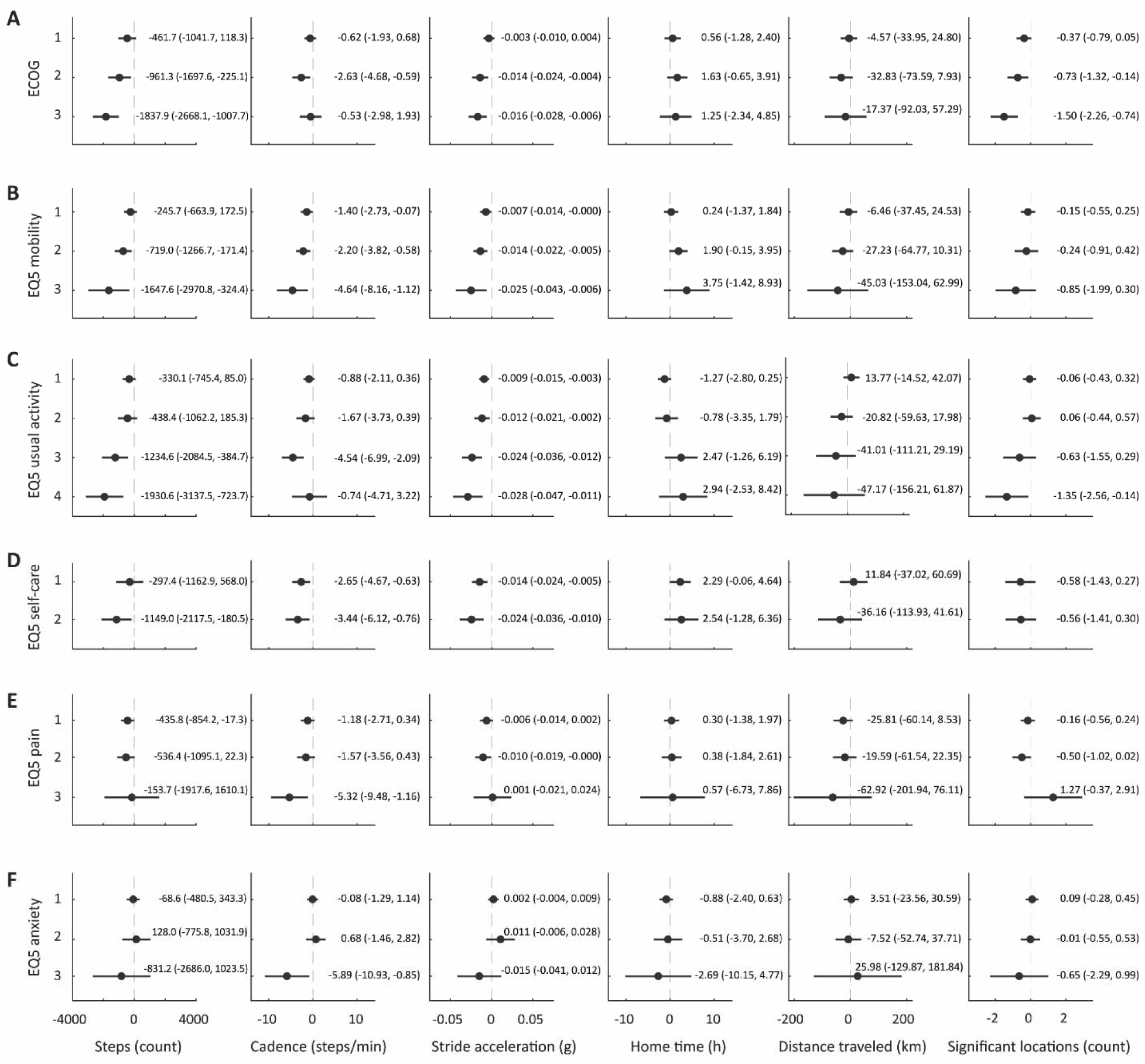
Forest plot of estimated regression coefficients with 95% confidence intervals for ECOG PS (panel A) and EQ-5D-5L items on mobility (B), usual activity (C), self-care (D), pain (E), and anxiety (F).

We also observed statistically significant associations between participants’ self-reported difficulty walking (**Figure 3, panel B**) and smartphone-based measures of participants’ step counts, cadence, and stride acceleration, such that participants who had higher EQ-5D-5L mobility scores and reported more difficulty walking had worse measures of gait-related physical activity. Participants with higher scores were also more likely to spend more time at home, travel shorter distances, and visit fewer significant locations.

We observed similar associations between participants’ self-reported ability to perform usual daily activities and smartphone-based measures of step counts and stride acceleration and inverse associations with mobility measures, although the latter results were not statistically significant (**Figure 3, panel C**). Similarly, we observed trends in decreased step-counts in participants who had more difficulty caring for themselves, compared with those who had no difficulty (**Figure 3, panel D**).

We observed weaker associations between participant-reported pain levels and smartphone-based measures of physical activity and mobility (**Figure 3, panel E**), as well as associations between anxiety symptoms and smartphone-based measures (**Figure 3, panel F)**.

The analysis examining associations between PROMIS PF 6b T-scores and digital measures of physical activity demonstrated statistically significant relationships, particularly for step counts and stride acceleration (**Table 2**). In these analyses, a one-point increase in PROMIS T-score was associated with an average of 72.6 more daily steps (*p*<0.001), 0.13 steps per minute in cadence (*p*=0.03) and 0.001 g faster stride acceleration (*p*<0.001), reflecting better physical function compared with lower PROMIS PF scores. In contrast, GPS-derived mobility measures demonstrated weaker and non-significant associations with PROMIS PF.

**Table 2.**
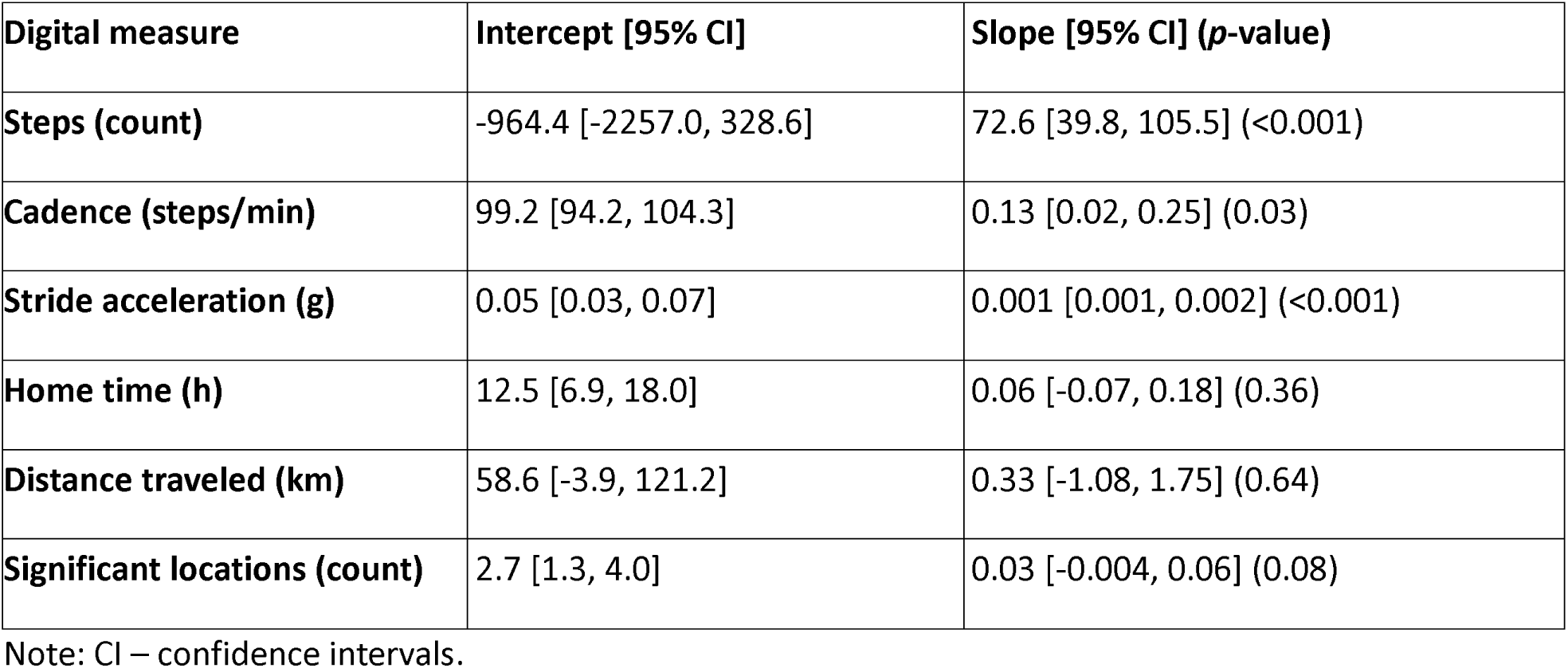
Estimated intercepts and slopes associated with one-point higher PROMIS PS 6b T-score.

### 3.4. Longitudinal trends in smartphone-based digital measures of physical function and mobility

Over the six-month study period, participants’ walking cadence decreased, although the absolute magnitude of this change was small (-0.23 steps/minute [-0.42, -0.03]; **Table 3**). We did not observe any statistically significant differences in other measures of physical activity or mobility.

**Table 3.** Average baseline and monthly change (per one month) of day-level outcomes derived from smartphone accelerometer and GPS data.

| Digital measure | Intercept [95% CI] | Slope [95% CI] (*p*-value) |
| --- | --- | --- |
| Steps (count) | 2,235.0 [1,846.0, 2,623.0] | -38.62 [-113.67, 36.42] (0.31) |
| Cadence (steps/min) | 105.5 [104.2, 106.7] | -0.23 [-0.42, -0.03] (0.02) |
| Stride acceleration (g) | 0.10 [0.09, 0.10] | -0.001 [-0.002, 0.001] (0.27) |
| Home time (h) | 14.9 [13.6, 16.3] | 0.04 [-0.31, 0.40] (0.81) |
| Distance traveled (km) | 79.0 [64.5, 93.5] | -2.93 [-6.98, 1.12] (0.16) |
| Significant locations (count) | 4.001 [3.7, 4.303] | -0.027 [-0.104, 0.050] (0.492) |

Note: CI – confidence intervals.

## 4. DISCUSSION

In this study we found that passively collected data from participants’ personal smartphones were strongly associated with key oncologic outcomes, including performance status, health-related quality of life, and physical functioning. Several gait-related measures, including daily step counts and stride acceleration, were particularly meaningful. For example, we found that participants with an ECOG performance of 3 (capable of only limited selfcare; confined to bed or chair more than 50% of waking hours) walked 1,837 fewer steps daily than participant with an ECOG of 0. Similarly, participants with better self-reported physical function had greater daily step counts, increased step cadence, and greater stride acceleration, compared with those who reported worse physical functioning. Together, these findings suggest that smartphone-based measures of physical activity may be a non-invasive, low-burden strategy for monitoring patients’ performance status and physical functioning objectively.

Prior research has demonstrated that oncologists’ estimates of patients’ performance status and physical functioning are subjective, only moderately correlated with one another[37,38], and weakly correlated with patient self-reports[39–41]. To address this gap, multiple researchers have tested whether commercial wearables can provide more objective data, demonstrating that commercial wearables can predict individuals with advanced cancers’ physical function[42], symptom burden, and risk of hospitalization or death[12,14,43–47]. While promising, commercial wearables are expensive and require patients to consistently wear, charge, and sync the devices[48,49]. In prior studies, adherence to wearables among individuals with cancer has varied widely based upon the context, ranging from 17% to 100%[48,50]. In contrast, smartphones have been widely adopted, have the capacity for passive data collection, and can be used to detect temporal changes in digital health biomarkers[51,52].

Indeed, a recent longitudinal study of cancer patients’ adherence to daily symptom surveys, commercial wearables, and/or smartphone data collection demonstrated that adherence was highest for smartphone data collection[49]. In that study, approximately 73% of participants contributed at least 8 hours of valid data. Here, we found that about three quarters of participants with advanced cancer contributed >16 hours of valid data, and the data quality did not differ with participants’ reported smartphone use, suggesting robust data collection regardless of overall phone usage patterns. The ubiquity of smartphones also enhances inclusivity, making this approach accessible to patients across diverse income levels and ethnic backgrounds[53,54], compared with commercial wearables.

In this study, smartphone-based mobility-related metrics provided further insights. For example, patients with worse EQ-5D-5L mobility and usual activity scores tended to spend more time at home, underscoring the behavioral consequences of limited mobility. Similarly, participants with worse ECOG PS and EQ-5D-5L scores traveled shorter distances and visited fewer significant locations, relative to participants with better performance status and health-related quality of life, although these associations were weaker compared to gait metrics. These findings suggest that while mobility measures may be less directly sensitive to physical function, they may capture distinct behavioral patterns associated with reduced activity levels and social engagement.

Despite these strengths, our study had several limitations. Although our data quality was comparable to other studies, nearly 25% of participant data was excluded due to technological and/or user issues limiting data quality (e.g., old operating systems, app uninstallation, sensor deactivation). Second, while gait metrics showed strong associations with PROMs, mobility measures demonstrated weaker associations. These findings indicate that mobility metrics may capture distinct behavioral dimensions, such as time spent in familiar environments or reduced social engagement, rather than direct physical impairments. Future research should explore how these mobility metrics can complement gait measures in providing a holistic view of patient health. In addition, participants were recruited from a single center. Further validation of these findings in more diverse cancer populations, including those with severe mobility impairments and those with different cancers or undergoing different treatments, is needed to ensure broad applicability. Expanding the scope of digital metrics to include social, cognitive, and emotional domains could also provide a more comprehensive picture of patient well-being.

In this study, we demonstrated that smartphone-based metrics, including both gait and mobility measures, offer a scalable, non-intrusive, and low-burden strategy for remote monitoring of physical function, health-related quality of life, and performance status in real-world settings for individuals with advanced cancer. While gait-related measures provide direct indicators of participants’ physical function and performance status, mobility metrics may capture the broader behavioral and environmental contexts of patient activity. Together, these metrics complement PROMs, offering oncologists actionable insights into patients’ activity levels and performance status. Addressing challenges related to data missingness and adherence will be essential for fully realizing the potential of this approach. By integrating these metrics into routine care, clinicians could improve the precision of performance assessments, enabling more timely and personalized care.

## ACKNOWLEDGMENTS

We would like to thank the patients in the HOPE study for their dedication to data collection.

## DECLARATIONS

### Data availability statement

The de-identified data are available upon reasonable request.

### Ethics approval statement

The study was approved by the Dana-Farber/Harvard Cancer Center Institutional Review Board.

### Patient consent statement

All participants provided written informed consent. ***Permission to reproduce materials from other sources:*** Not applicable.

### Clinical trial registration

The study was registered in clincialtrials.gov (NCT03022032).

## AUTHORS CONTRIBUTION

MS – Study concept and design, method development, data collection, data processing, data analysis, figure and table preparation, manuscript drafting, and final approval

NLK – Study concept and design, data interpretation, critical review of manuscript, and final approval.

SMS – Data collection, critical review of manuscript, and final approval.

UAM – Data collection, critical review of manuscript, and final approval.

NH – Data collection, critical review of manuscript, and final approval.

SC – Data collection, critical review of manuscript, and final approval.

JPO – Study concept and design, method development, data collection, data analysis, critical review of manuscript, scientific supervision.

AAW – Study concept and design, data collection, data analysis, critical review of manuscript, scientific supervision.

